# Energy-loss index does not improve risk prediction in aortic stenosis compared to conventional aortic valve area assessment

**DOI:** 10.1101/2023.03.08.23287015

**Authors:** Thomas Lindow, David Playford, Geoff Strange, Rebecca Kozor, Martin Ugander, the NEDA Contributing Sites, Fremantle, Adelaide, Sydney, Melbourne, Brisbane, and Casuarina, Australia

## Abstract

**Background:** Evidence of improved risk assessment in aortic stenosis (AS) by using energy-loss index (ELI) instead of aortic valve area indexed to body surface area (AVAi) is scarce, and positive results have been driven by aortic valve replacement. We aimed to evaluate the prognostic performance of ELI and AVAi in a head-to-head comparison using large-scale, real-world data.

**Methods:** In the multi-center, mortality-data linked National Echocardiography Database of Australia (NEDA), patients with AS and requisite ascending aortic area measurements were identified. The prognostic value of AVAi and ELI, respectively, was analyzed using Cox regression and the C statistic.

**Results:** In patients with mild AS (n=3,179), moderate AS (n=4,194), and severe AS (n=3,120), there were 4,229 deaths of which 2,359 were reported as cardiovascular deaths (median [interquartile range] follow-up 2.5 [1.1–4.5] years]. Decreasing AVAi was associated with increased cardiovascular mortality (hazard ratio [95% confidence interval] 1.18 [1.16– 1.20] per 0.1 cm^2^/m^2^ downward increment]. Prognostic performance for 5-year mortality did not improve by using ELI instead of AVAi (identical C statistics 0.626 [0.612–0.640]), and the relative performance did not change when analyzing 1-year cardiovascular mortality, or all-cause mortality.

**Conclusion:** ELI was not associated with improved prognostic performance compared to AVAi in echocardiographic assessment of AS using large-scale, real-world clinical data. AVAi remains a relevant measure for risk prediction in AS, providing information on incremental risk with decreasing area.

## Clinical perspective

Overestimation of pressure gradients in aortic stenosis (AS) can be explained by the pressure recovery phenomenon. By taking this pressure recovery into account, the true net pressure gradient across the valve can be appreciated. While catheter-based gradients are measured beyond the stenosis and after pressure recovery, Doppler-derived gradients are based on velocity measures within the vena contracta, and any pressure recovery distal to this site will result in overestimation of the true net gradient. This can be accounted for using energy-loss index (ELI). Evidence of improved risk assessment by ELI is scarce. We aimed to evaluate the prognostic performance of both ELI and aortic valve area (AVA) indexed to body-surface area regarding cardiovascular death using large-scale, real-world data; the multi-center, mortality-data linked National Echocardiography Database of Australia (NEDA). We included 10,443 patients with mild, moderate or severe AS. There were 2359 cardiovascular deaths during a median follow-up of 2.5 years. Both AVA and ELI were associated with increased risk of cardiovascular death, but there was no difference in c statistics, i.e. ELI was not associated with improved prognostic performance compared to AVA. Possibly, this can be explained either by increased measurement errors using ELI, or that ELI does not fully account for the complexity in pressure recovery, or that other factors than the pressure gradient contribute to mortality.

## Background

Echocardiography is the non-invasive method of choice in aortic stenosis (AS) assessment.^1^ Doppler ultrasound is used to estimate the pressure gradient across the aortic valve,^1–3^ and although good agreement can be achieved between Doppler-derived pressure gradients and catheter gradients, overestimation of pressure gradients by echocardiography is not uncommon.^4, 5^ This can be explained by the pressure recovery phenomenon.^4–7^ When blood is accelerating through a stenotic aortic valve opening, static pressure energy is converted into kinetic energy. Upon entering the ascending aorta, blood flow decelerates and some of the kinetic energy is dissipated into heat and some is recovered as static pressure energy, depending on the amount of turbulent flow. By taking this recovery of pressure into account, the true net pressure gradient across the stenotic valve can be appreciated. While catheter-based pressure gradients are measured distantly from the stenosis where pressure recovery already will have occurred, Doppler-derived pressure gradients are based on velocity measures within the *vena contracta*, and any pressure recovery distal to this site will thus result in overestimation of the true net gradient. The pressure recovery phenomenon has been demonstrated to occur in several experimental studies, both in prosthetic and native aortic valves ^4, 6–10^, and suggestions have been made on how to adjust for this in echocardiography.^10^ Since pressure recovery increases with increasing aortic valve area (AVA) and decreasing aortic area (AoA), a pressure recovery adjusted AVA, also known as the energy-loss index (ELI), can be defined with a simple formula incorporating AVA, AoA and body surface area.^10^ Moving from smaller experimental studies to larger epidemiological studies, attempts have been made to prove that pressure recovery is a clinically relevant parameter in the risk assessment of patients with AS.^11–14^ However, evidence of improved risk assessment by using ELI instead of unadjusted AVA is scarce, and results have either been driven by aortic valve replacement numbers^10, 13^ or flawed by not considering pre-existing differences between patients that were reclassified after pressure recovery adjustment to those that were not^12, 14^ Despite previous attempts, it is still not known whether pressure recovery adjustment improves risk stratification of AS patients. Therefore, we aimed to evaluate the prognostic value of both AVAi and ELI in a head-to-head comparison using large-scale, real-world data. The primary outcome was the prediction of CV mortality, and the secondary outcome evaluated all-cause mortality. We also aimed to evaluate the association between AVA and ELI, respectively, and the hemodynamic consequences of aortic stenosis, including left ventricular hypertrophy, left atrial enlargement, increased pulmonary artery pressure, and increased filling pressures.

## Methods

The National Echo Database Australia (NEDA) is a large, observational registry including individual echocardiographic data on a retrospective and prospective basis from participating centers throughout Australia.^15^ Currently, >600,000 subjects are included in the registry. Typically, included subjects have been referred by a primary care physician for echocardiography in the investigation of suspected heart disease or as part of routine management of known CV disease. In accordance with the structure of the Australian health care system, minimal referral bias applies.

Data have been cross-linked to the Australian National Death Index^16^ to obtain survival status for each subject until study census date (21 May 2019). In consistency with previous NEDA analyses,^17–19^ causes of death were categorized according to ICD-10 and a primary code within I.00 – I.99 was considered as a CV-related death.

The NEDA database has been registered in the Australian New Zeeland Clinical Trials Registry [ACTRN12617001387314]. Ethical approval has been obtained from all relevant Human Research Ethics Committees.

### Echocardiography

All measurements from investigations performed at any of the NEDA participating centers from 1 Jan 2000 to 21 May 2019, and accompanying data on sex, date of birth, date of examination, weight, and height, were transferred to a central database through an automated process for data extraction. All subjects included in the NEDA database are given a unique ID linked to their echocardiographic investigations. For this study, only the last echocardiogram for each eligible subject was included.

For this study, adult patients with native valve aortic stenosis (peak aortic velocity >2.5 m/s and/or AV mean gradient ≥10 mmHg)^1^ were eligible for inclusion. Patients were excluded if follow-up data or data on aortic peak velocity were missing, in case of previous AVR, or if severe/moderate to severe aortic/mitral regurgitation were present. Finally, patients were excluded if data necessary to perform pressure recovery adjustment calculations (AVA, AoA) were missing. A flow-chart of patient inclusion and exclusion is presented in Figure 1.

**Figure 1.**
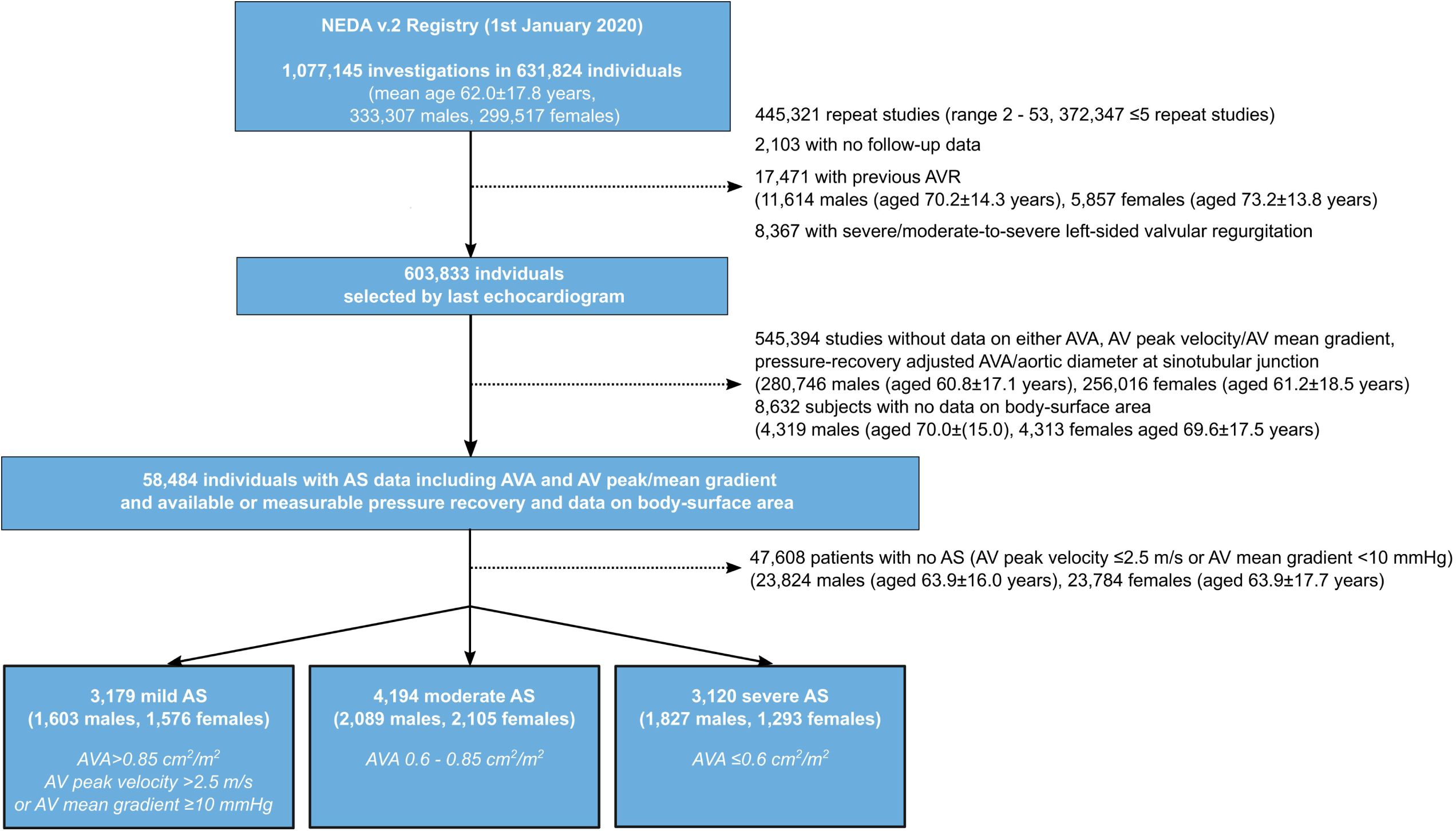
Flow chart of patient inclusion and exclusion.

Patients were graded based on AVAi according to the following criteria: severe AS: ≤0.6 cm^2^; moderate AS: 0.6 – 0.85 cm^2^; and mild AS: >0.85 cm^2^ and an aortic peak velocity >2.5 m/s and/or a mean gradient ≥10 mmHg. Severe and moderate AS included patients with both high- and low-gradient AS, and their proportions are described in Figure 1.

ELI was calculated in all patients according to the following formula:^10^

*Energy-loss index, ELI = (AVA × AoA) / (AoA – AVA)/body surface area*

*in which*

*aortic valve area, AVA = r_LVOT_*^2^ *× VTI_LVOT/_ VTI_AV_*

*and*

*Aortic area, AoA = π × r* ^2^

VTI_LVOT_ denotes the velocity time integral at the left ventricular outflow tract, VTI_AV_ denotes the velocity time integral at the aortic valve, and r_stj_ denotes the aortic radius at the sinotubular junction. Left ventricular (LV) mass was calculated according to the Cube formula: LV mass (g) = 0.8 × 1.04 ([IVS + LVID + PW]^3^ – LVID^3^) + 0.6. Increased LVM was defined as any value above the sex-specific upper limit of normal.^20^ Left atrial volume is presented indexed to body surface area (LAVi) and increased LAVi was defined as LAVi >34 ml/m^2^.^20^ Increased right ventricular systolic pressure gradient was defined as tricuspid regurgitation peak velocity above 2.8 m/s. E/e’ ratio was considered elevated when >14.^21^

## Statistical analysis

Statistical analysis was performed using R v. 3.5.3 (R Core Team (2021). R: A language and environment for statistical computing. R Foundation for Statistical Computing, Vienna, Austria (https://www.R-project.org/), example packages: Survival v. 3.1-12, OptimalCutpoints v.1.1.4, MASS v. 7.3.53.1). Continuous variables were described using mean and standard deviation (SD). Proportional differences between groups were assessed using the χ2 test. Comparison of group means were performed using Student’s t test. The association between AVAi and mortality, and ELI and mortality, respectively, was analyzed using multivariable Cox proportional hazard regression models adjusted for age, sex and AV peak velocity. Hazard ratios (HR) are presented with 95% confidence intervals (CI) with increased risk by decreasing AVA per 0.1 cm^2^/m^2^ increments presented as HR above 1. Since risk of death was expected to increase substantially at lower values of either AVAi or ELI, natural cubic splines were used to characterize the risk associated the with decreasing AVAi and ELI values, respectively, using four knots placed at the 5^th^, 35^th^, 65^th^ and 95^th^ percentiles. The assumption of proportional hazards was confirmed using Schoenfeld’s residuals. Comparison of predictive strength of the models was performed by reporting the C statistic (concordance), and by describing the diagnostic accuracy at optimal cut-offs. Based on the highest Youden’s index, optimal cutoffs for 5- and 1-year CV mortality were decided for both AVAi and ELI, respectively.

Reclassification was considered to occur if a patient received a lower grade of AS severity using ELI compared to AVAi, e.g. severe by AVAi but moderate by ELI (>0.6 cm^2^/m^2^ but <0.85 cm^2^/m^2^), as per previous studies on pressure recovery adjustment of AS grade ^11, 12, 14^. Cox regression was used to determine whether reclassification from either severe to moderate AS, or from moderate to mild was associated with improved survival. This was determined by using reclassification as a binary variable and 5-year CV death as the outcome variable. To determine whether the association between reclassification and survival was determined by differences in AVAi between those that were reclassified and those that were not, the same analysis was performed with adjustment for AVAi. This was done to evaluate whether a potentially positive effect of reclassification was confounded by how close the unadjusted area measurement was to the specific threshold.

In addition, the following sensitivity analyses were performed: a) including only patients with an aortic diameter ≤3.0 cm (since reclassification is more common in patients with smaller aortic diameter), b) including only patients with either moderate or severe AS but excluding patients with low-gradient AS, and c) including only patients with similar stroke volume (60 – 80 ml, since pressure recovery depends not only on aortic area but also on flow rate^10^). Sensitivity analyses were performed for the Cox regression analysis and reported with C statistics with 95% confidence intervals (95%CI).

The correlation between AVAi or ELI and LV mass, left atrial volume, right systolic ventricular pressure gradient, and E/e’ ratio was assessed using Pearson’s r, and multivariable analysis was performed using linear regression with backward elimination at p≥0.1, and logistic regression for binary outcomes of increased LV mass, left atrial enlargement, elevated, E/e’ and right systolic ventricular pressure gradient based on commonly used reference values as described above. Statistical significance was accepted at the level of p<0.05 (two-sided).

## Results

In total, 10,493 patients were included in the study (73.9±14.5 years, 47% females). Baseline characteristics are presented in Table 1. According to AVAi, 3,179 patients had mild AS, 4,194 patients had moderate AS and 3,120 patients severe AS (Fig. 1).

**Table 1.**
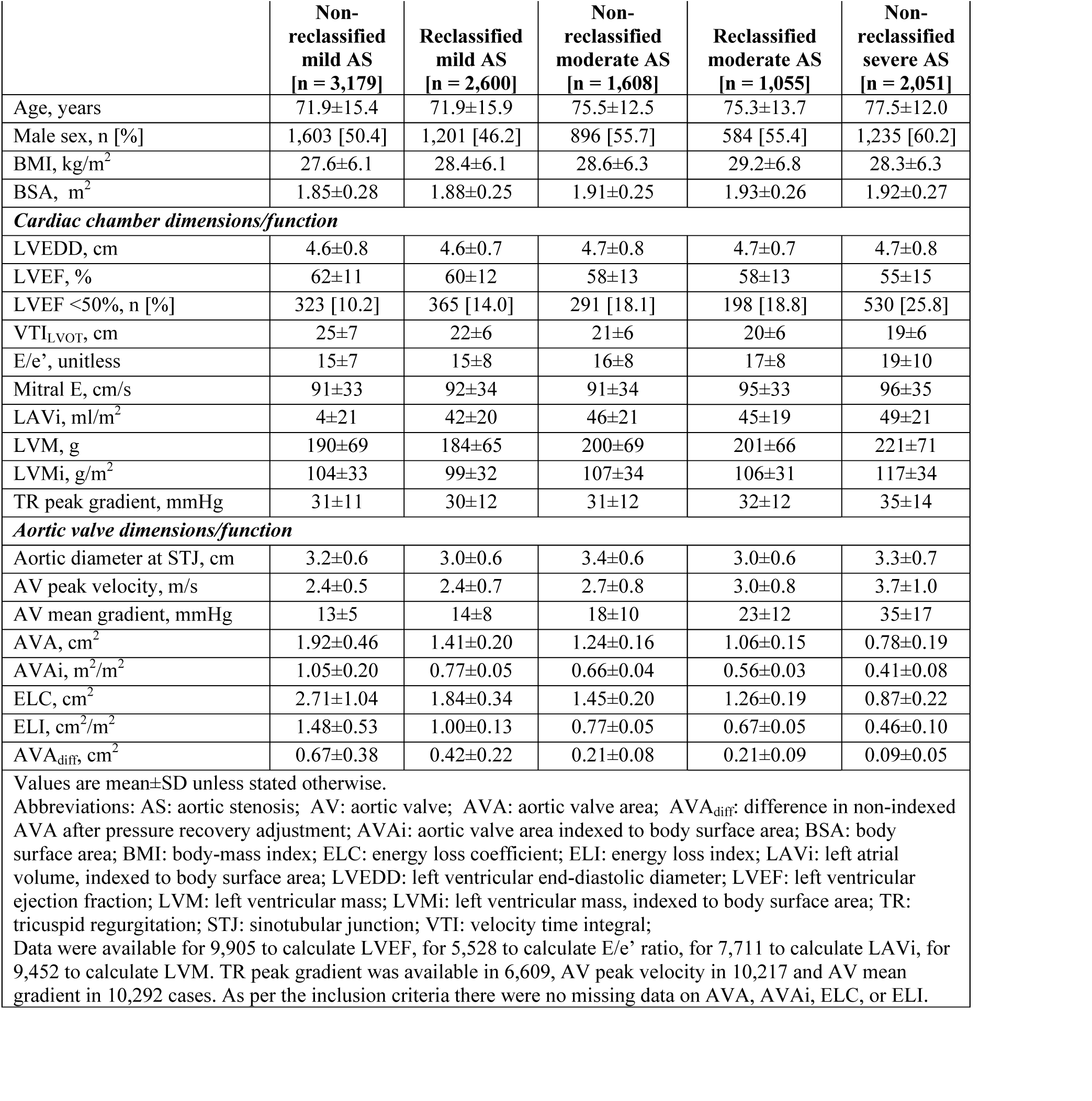
Baseline characteristics [n = 10,493)

During a median follow-up of 2.5 years (interquartile range 1.1–4.5 years), 4,229 patients (40%) died, of whom 2,359/4,229 (56%) due to a primary CV cause. One- and 5-year mortality rates by reclassified or non-reclassified AS grades are presented in Table 2.

**Table 2.** One- and five-year mortality stratified by reclassified and non-reclassified aortic stenosis grades

|  |  | Mild AS | Reclassified mild AS | Non-reclassified moderate AS | Reclassified moderate AS | Non-reclassified severe AS | p |
| --- | --- | --- | --- | --- | --- | --- | --- |
| 1-year mortality* | All-cause | 385 (13.0) | 314 (13.0) | 259 (17.8) | 200 (20.1) | 577 (30.6) | <0.001 |
|  | CV | 211 (7.1) | 193 (8.0) | 156 (10.8) | 148 (14.9) | 406 (21.5) | <0.001 |
| 5-year mortality** | All-cause | 545 (40.1) | 462 (44.1) | 336 (55.7) | 240 (55.7) | 646 (66.1) | <0.001 |
|  | CV | 335 (24.7) | 282 (26.9) | 225 (37.3) | 172 (39.9) | 480 (49.1) | <0.001 |

Data are presented as number (%).
\* In patients with at least 1-year possible follow-up
\*\* In patients with at least 5-years possible follow-up (n=4,416)
Abbreviations: AS: aortic stenosis; CV: cardiovascular

### Reclassification due to pressure recovery adjustment

After pressure recovery adjustment, 34% of patients were reclassified from severe to moderate (n=1,055) or mild (n=14), and 45% from moderate to mild (n=2,586). Reclassification from severe to moderate was most frequent in patients with small aortic diameter (60.1% in patients with aortic diameter <2.5 cm, 41.8% in patients with aortic diameter 2.5 – 3.0 cm), but was common also among those with larger diameter (22.8% in patients with aortic diameter 3.5 – 4.0 cm, 13.1% in patients with aortic diameter 4.0 – 4.5 cm). The aortic diameter was smaller in female patients than in male (3.0±0.6 vs. 3.3±0.6 cm, p<0.001). Reclassification occurred more often in female than in male patients (43.1% vs. 27.7%, p<0.001).

Both 5- and 1-year, CV and all-cause mortality was highest for patients with severe AS, and declined for each reduction in grade of severity (Table 2). Patients that were reclassified from severe to moderate had lower mortality rates than those with non-reclassified severe AS (p<0.001), and patients that were reclassified from moderate to mild, had lower mortality rates than those with non-reclassified moderate AS (p<0.001). Compared to those that were reclassified to moderate AS, patients with non-reclassified severe AS had higher AV mean gradient (35±17 vs. 24±12 mmHg, p<0.001), higher AV peak velocity (3.7±0.9 vs. 3.0±0.8 m/s, p<0.001), and lower AVAi (0.41±0.08 vs. 0.56±0.03 cm^2^, p<0.001). Similarly, compared to those that were reclassified to mild AS, patients with non-reclassified moderate AS had higher AV mean gradient (18±10 vs. 14±8 mmHg, p<0.001), higher AV peak velocity (2.7±0.7 vs. 2.4±0.7 m/s, p<0.001), and lower AVAi (0.66±0.04 vs. 0.77±0.05 cm^2^, p<0.001).

In patients with severe AS, reclassification to moderate AS was associated with reduced risk of death (death within 5 years, HR 0.73 [0.63–0.84] but not if AVAi was taken into account (HR 1.04 [0.86–1.26]). Similarly, in patients with moderate AS, reclassification to mild AS was associated with reduced risk of death (death within 5 years, HR 0.73 [0.63–0.84]) but not once AVAi was added to the model (HR 0.92 [0.74 – 1.14]).

### Prognostic and hemodynamic consequences of pressure recovery aortic valve area adjustment

Decreasing AVAi and ELI were both associated with increased mortality, both for all-cause mortality and CV mortality (Table 3), and the risk increased exponentially with decreasing AVAi and ELI, respectively (Fig. 2). No difference was found in the C statistic between AVAi and ELI measures, neither for CV mortality nor for all-cause mortality (Table 3), and similar results were found in the sensitivity analyses (Table S1).

**Table 3.** Hazard ratios (HR, unadjusted and adjusted for age, sex and AV peak velocity) and C statistics for aortic valve area indexed to body surface area and energy-loss index in the prediction of cardiovascular mortality and all-cause mortality at 5 years.

|  | **Cardiovascular mortality** |  |  |  |
| --- | --- | --- | --- | --- |
|  | Absolute estimate |  | Adjusted for age, sex and AV peak velocity |  |
|  | HR [95% CI] | C statistic | HR [95% CI] | C statistic |
| AVAi, per 0.1 cm<sup>2</sup>/m<sup>2</sup> decrement | 1.18 [1.16–1.20] | 0.626 [0.612–0.640] | 1.14 [1.12–1.16] | 0.711 [0.699–0.723] |
| ELI, per 0.1 cm<sup>2</sup>/m<sup>2</sup> decrement | 1.10 [1.09–1.12] | 0.626 [0.612–0.640] | 1.07 [1.06–1.09] | 0.707 [0.695–0.719] |
|  | **All-cause mortality** |  |  |  |
|  | Absolute estimate |  | Adjusted for age, sex and AV peak velocity |  |
|  | HR [95% CI] | C statistic | HR [95% CI] | C statistic |
| AVAi, per 0.1 cm<sup>2</sup>/m<sup>2</sup> decrement | 1.12 [1.11–1.14] | 0.593 [0.583–0.601] | 1.09 [1.07–1.10] | 0.691 [0.683–0.699] |
| ELI, per 0.1 cm<sup>2</sup>/m<sup>2</sup> decrement | 1.07 [1.06–1.08] | 0.597 [0.587–0.607] | 1.05 [1.04–1.05] | 0.689 [0.681–0.697] |
| Abbreviations: AVAi: aortic valve area indexed to body surface area; CI: confidence interval; ELI: energy loss index; HR: hazard ratio |  |  |  |  |

Optimal cutoffs for either 5-year or 1-year cardiovascular mortality were higher for AVA_adj_ than for AVA, but there was no difference in accuracy (Table 4).

**Figure 2.**
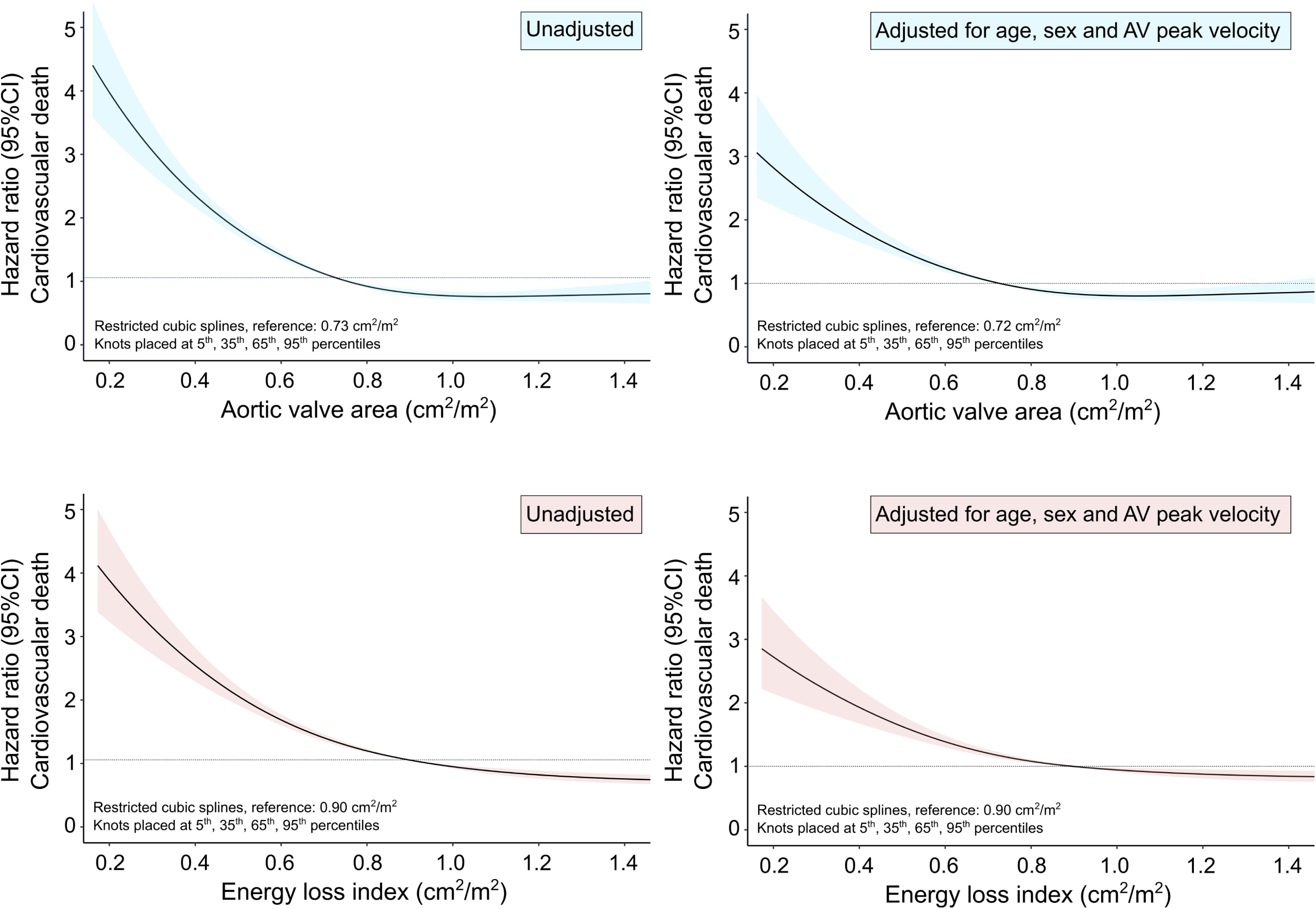
Impact of decreasing aortic valve area indexed by body surface area (AVAi) (upper panels) and energy loss index (ELI) (lower panels) on the risk of cardiovascular (CV) death. The hazard ratio (with 95% confidence intervals (CI)) for CV mortality was calculated using Cox regression and modelled with natural cubic splines with four knots, unadjusted (left panels), and adjusted for age, sex, and aortic valve (AV) peak velocity (right panels). Both AVAi and ELI showed exponentially increasing risk with decreasing area, but at different absolute values due to differences in magnitude.

**Table 4.** Optimal cutoffs based on the highest Youden’s index for body surface indexed aortic valve area and energy loss index for either 5-year or 1-year cardiovascular mortality

|  |  |  |  |
| --- | --- | --- | --- |
|  | **5-year cardiovascular mortality** |  |  |
|  | Optimal cut-off | Sensitivity [%] | Specificity [%] |
| AVAi, cm<sup>2</sup>/m<sup>2</sup> | 0.63 | 57.3 [54.3 – 60.2] | 66.7 [65.8 – 67.7] |
| ELI, cm<sup>2</sup>/m<sup>2</sup> | 0.75 | 55.7 [52.7 – 58.7] | 67.5 [66.6 – 68.5] |
|  | **1-year cardiovascular mortality** |  |  |
|  | Optimal cut-off | Sensitivity [%] | Specificity [%] |
| AVAi, cm<sup>2</sup>/m<sup>2</sup> | 0.63 | 57.3 [54.3 – 60.2] | 66.7 [65.8 – 67.7] |
| ELI, cm<sup>2</sup>/m<sup>2</sup> | 0.75 | 55.7 [52.7 – 58.7] | 67.5 [66.6 – 68.5] |
| Abbreviations: AVAi: aortic valve area indexed to body surface area; ELI: energy loss index |  |  |  |

For male patients, the C statistic was 0.629 [0.613–0.645] for AVAi in the prediction of CV death within 5 years, and 0.624 [0.608–0.640] for ELI. For female patients, the C statistic was 0.647 (0.631–0.662) for AVA in the prediction of CV death within 5 years, and 0.618 (0.598–0.638] for ELI.

AVAi and ELI were both significantly but weakly correlated with LV mass, LAVi, E/e’ ratio, and tricuspid regurgitation peak gradient. Both AV peak velocity and AV mean gradient showed stronger correlation with LV mass compared to both AVA measures (Table 5). In univariable analysis, both AVAi and ELI were associated with increased risk of increased LAVi, LV mass, tricuspid regurgitation peak gradient, and elevated E/e’ ratio. However, these associations did not persist in multivariable analysis (Table S2).

**Table 5.** Correlation coefficients and 95% confidence intervals for aortic valve area indexed to body surface area, energy loss index, aortic valve peak velocity and aortic valve mean gradient.

|  | Left ventricular mass<br>index | Left atrial volume<br>index | E/e' ratio | Tricuspid regurgitant<br>peak gradient |
| --- | --- | --- | --- | --- |
| AVA | -0.08 [-0.10 – -0.06] | -0.05 [-0.07 – -0.03] | -0.13 [-0.15 – -0.11] | -0.10 [-0.12 – -0.08] |
| ELI | -0.11 [-0.12 – -0.09] | -0.07 [-0.09 – -0.04] | -0.12 [-0.15 – -0.10] | -0.10 [-0.12 – -0.07] |
| AV peak velocity | 0.24 [0.22 – 0.26] | 0.10 [0.08 – 0.12] | 0.13 [0.10 – 0.16] | 0.15 [0.12 – 0.17] |
| AV mean gradient | 0.23 [0.22 – 0.25] | 0.10 [0.07 – 0.12] | 0.12 [0.10 – 0.17] | 0.13 [0.11 – 0.15] |

Abbreviations: AV: aortic valve; AVA: aortic valve area indexed to body surface area; ELI: energy loss index.

## Discussion

The main finding of the study is that although both decreasing AVAi and ELI were associated with increased risk of CV death, there was no incremental prognostic value in using ELI instead of unadjusted AVAi. This is the largest study evaluating the value of ELI assessment for the prediction of CV and all-cause mortality in AS patients.

ELI instead of AVAi did not improve predictions across the entire spectrum of AS, and ELI did not perform better than AVAi in predicting hemodynamic consequences of AS. These findings contrast previous studies on the prognostic value of pressure recovery adjustment of AS severity.^10, 12–14^ After showing *in vitro* that the energy loss could be accounted for using information on AVA and aortic area only, 138 patients with moderate or severe AS with a combined endpoint of death or AVR were evaluated over 8 months follow-up.^10^ In that study, ELI was the only significant aortic valve measure associated with survival in multivariable analysis. However, the study was small, and both AVAi and ELI were included in the same model. Consequently, multi-collinearity may have led to unreliable estimates. In a substudy of the Simvastatin and Ezetimibe in Aortic Stenosis (SEAS) study, in which patients with AS were randomized to lipid-lowering treatment or placebo, the prognostic value of ELI was evaluated in 1,563 patients with a median follow-up of 4.3 years for the combined outcome of AVR, hospitalization for heart failure resulting from AS progression, and CV death. The use of ELI was associated with a higher rate of the combined endpoint, mainly driven by AVR. Importantly, AVR as an endpoint to elucidate whether ELI adds incremental information is questionable given that, in these retrospective studies, the interventional decision was unlikely based on ELI. Perhaps most importantly, pressure recovery adjustment of AVA will, by mathematical definition, always result in larger estimations of AVA. Consequently, patients with low ELI will, by mathematical definition, have a lower AVAi (Fig. 3). At the same threshold for AVAi and ELI, it is thus expected that a higher rate of AVR will be performed in the ELI group since their adjusted AVAi is lower. Thus, AVAi and ELI cannot be compared using the same threshold for intervention. In line with this, the current study shows that the optimal cutoffs in prediction of 5-as well as 1-year mortality is consistently higher for ELI than for AVAi, but without improvement in accuracy. In the current study, the optimal cutoff for ELI (0.75 cm^2^/m^2^) was higher than that found by previous authors (0.52 cm^2^/m^2^ ^10^ and 0.60 cm^2^/m^2^ ^11^). Of note, for both AVA measures in the current study, sensitivity and specificity value were low, indicating that several other factors are likely in play in the prediction of mortality in AS patients. Also, it deserves to be mentioned that risk increases as a continuum along the AS severity spectrum in a manner that is not accurately reflected by a dichotomous cutoff, and cutoff values derived from an observational study do not offer immediate guidance on which value should be used to decide the most appropriate interventional strategy.

**Figure 3.**
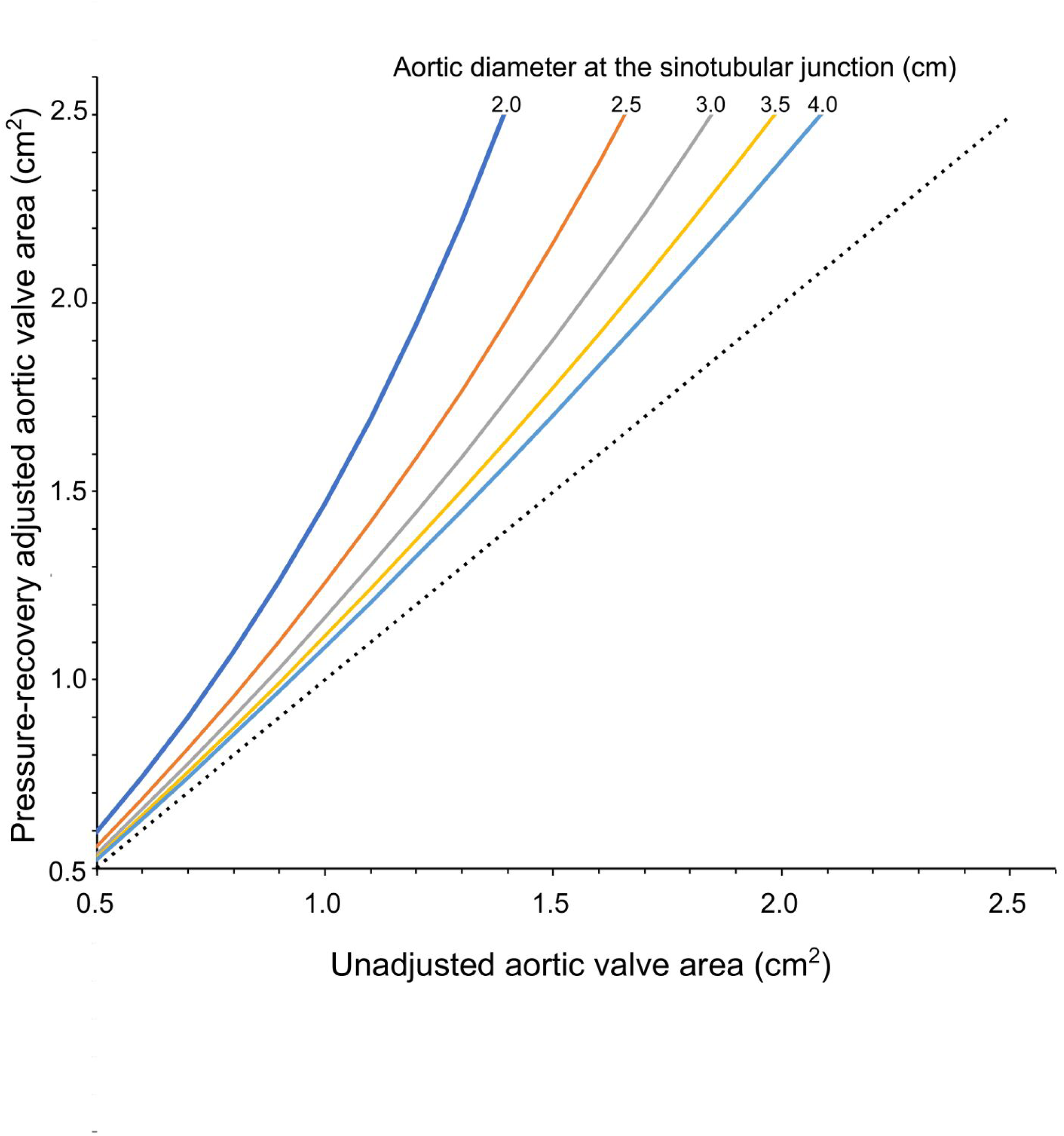
The relationship between non-adjusted aortic valve area and pressure recovery adjusted aortic valve areas for different aortic diameters. Note that the pressure recovery adjusted aortic valve area will, by mathematical definition, always be larger than the un-adjusted measure.

The observed risk reduction in reclassified patients disappear after adjusting for pre-existing differences in AVAi. Thus, the positive effects of reclassification are highly confounded by how close the unadjusted area measurement is to the threshold in question. At first glance, even in the current study, reclassification from severe to moderate, and from moderate to mild, seems to be associated with a reduction in risk, and such findings have been described in two prior studies.^12, 14^ One previous study aimed to determine the clinical implication of pressure recovery by studying the rate of AS severity grade reclassification due to pressure recovery adjustment, and comparing the risk between patients that were reclassified to those that were not.^12^ In that study, almost 25% of patients with severe AS could be reclassified to moderate AS after adjusting for pressure recovery and showed higher event-free survival rates compared to patients with AS who remained severe after pressure recovery adjustment.

However, the improved survival is expected, since those who remained classified as severe had lower AVA and higher AV mean gradient, and would therefore always, by definition, be at higher risk.^22^ In a recent study, improved survival among patients undergoing transcatheter aortic valve implantation was observed for patients that were reclassified after pressure recovery adjustment and multimodal imaging of the left ventricular outflow tract. These patients had lower AV mean gradient, as well as lower Agatson scores.^14^ Of note, the same principles of echocardiographically assessing AS severity before intervention apply regardless of whether the aortic valve intervention is surgical or using a transcatheter approach. Hence, there is no reason why pressure recovery adjustment would have different consequences for different interventional methods.

In agreement with our findings, in a small, observational study of asymptomatic patients with AS, ELI did not improve prediction of major cardiac events (including cardiac death, ventricular fibrillation, myocardial infarction, heart failure requiring admission, and aortic valve replacement). However, a small benefit was observed among those with severe AS.^23^ Our findings do not, per se, contradict the existence nor the effect of the pressure recovery phenomenon, which has been shown in several studies, both *in vitro* and *in vivo*.^4–10^ The lack of impact on hard endpoints such as mortality, could instead be explained either by a) that the use of additional measures in AS assessment increases the risk of measurement errors which could cloud the true impact, b) that the suggested correction formula does not account for the complexity in pressure recovery, or c) that other factors than the net pressure gradient contribute to mortality in this population.

The complexity of pressure recovery may not be fully appreciated in the “easy-to-use” equation provided by Garcia, et al.^10^ For instance, although flow rate is known to affect pressure recovery, the equation assumes similar flow rates by adjusting for body surface area.^10^ However, transaortic flow rates vary and has been shown to be associated with mortality in AS patients.^24–26^ In this study, we were unable to account for flow rate since ejection time was rarely recorded. Other approaches which to a greater extent directly measure the energy loss may prove to be more successful. Using cardiovascular magnetic resonance imaging (CMR), for example, the flow effects in AS can be assessed and thus the turbulent kinetic energy, which dominates the energy dissipation into heat, can be estimated.^27, 28^ However, the CMR techniques need further validation.^27, 28^

Other factors than the net pressure gradient are likely to contribute to mortality. Prior observations have shown that patients with moderate AS are at almost similarly high risk as those with severe AS, despite having lower gradients.^18, 29^ The current study found the risk to increase exponentially. This extends the knowledge from a previous study where it was observed that decreasing AVA in patients with severe AS (AVA ≤1.0 cm^2^) was incrementally associated with an increasing 5-year mortality.^30^

In the current study, increasing mortality with decreasing AVA was observed in spite of weak correlations to hemodynamic measures such as LV mass or surrogates of increased filling pressures. This is in agreement with the findings by Dweck, et al, who investigated the patterns of LV hypertrophy in patients with moderate and severe AS (AV peak velocity ≥3.0 m/s and AVA <1.5 cm^2^), and found that the degree of AS severity was unrelated to LV mass by CMR.^31^ Also, although heart failure in patients with severe AS is associated with a particularly poor prognosis, HF has multiple etiologies beyond LV pressure load due to valvular obstruction, e.g. hypertension, ischemic heart disease, infiltrative myocardial disease, which can occur at any grade of AS.^32, 33^

Our results are limited by the lack of important clinical information such as concomitant CV disease, medications, biomarkers, or future AVR. This does not have a sizable effect on the results regarding the primary aim of this study, i.e. to compare AVAi vs. ELI, since that comparison is made with the same data, and the same limitations thus are present for both AVA measures. However, we cannot conclude from our data whether ELI better discriminates symptomatic AS from asymptomatic AS. Also, since clinical management decisions have been made, likely to a very large extent at least, without knowledge of ELI, we do not know if the outcome would have been different if patients were managed according to ELI values instead of conventional measurements. However, this applies to all existing studies on the prognostic value of pressure recovery.^10–13, 23^ For this to be determined, a randomized clinical trial in which patients are managed based on either ELI or AVAi is needed, e.g. in patients with smaller aortic diameters in which the impact of pressure recovery is greater.

## Conclusion

In real-world clinical data, pressure recovery adjustment of aortic valve area was not associated with improved risk prediction in echocardiographic assessment of aortic stenosis compared to using aortic valve area alone. Despite this, aortic valve area is a relevant measure for risk prediction in AS patients, providing information on incremental risk with decreasing aortic valve area.

## Data Availability

The data that support the findings of this study are available from the corresponding author upon reasonable request

---

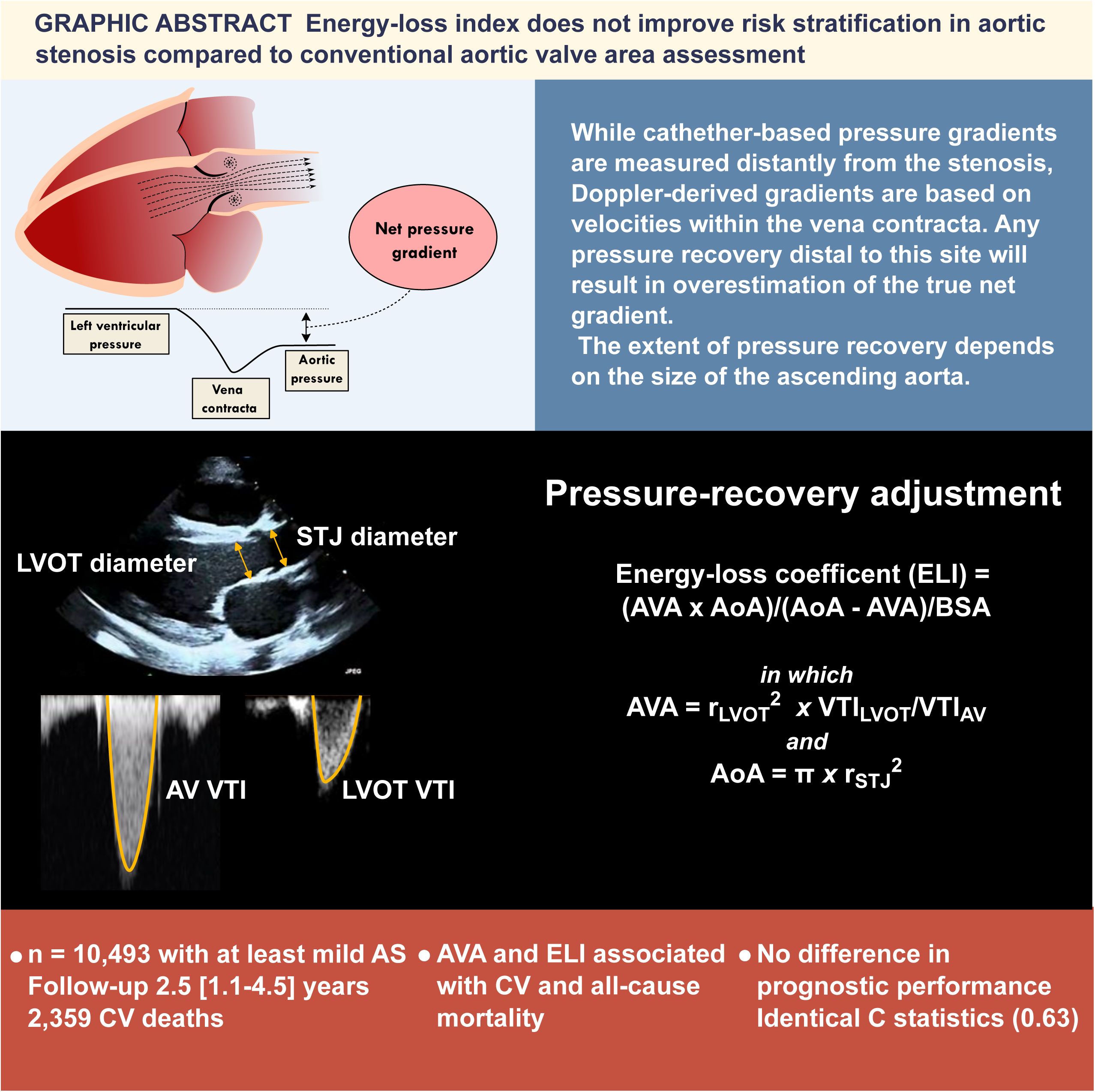

## References

1. Baumgartner H, Falk V, Bax JJ, De Bonis M, Hamm C, Holm PJ, et al. 2017 ESC/EACTS Guidelines for the management of valvular heart disease. Eur Heart J. 2017;38:2739–2791.

2. Hatle L, Angelsen BA and Tromsdal A. Non-invasive assessment of aortic stenosis by Doppler ultrasound. Br Heart J. 1980;43:284–92.

3. Vahanian A, Beyersdorf F, Praz F, Milojevic M, Baldus S, Bauersachs J, et al. 2021 ESC/EACTS Guidelines for the management of valvular heart disease. Eur Heart J. 2021.

4. Baumgartner H, Stefenelli T, Niederberger J, Schima H and Maurer G. “Overestimation” of catheter gradients by Doppler ultrasound in patients with aortic stenosis: a predictable manifestation of pressure recovery. J Am Coll Cardiol. 1999;33:1655–61.

5. Baumgartner H, Khan S, DeRobertis M, Czer L and Maurer G. Discrepancies between Doppler and catheter gradients in aortic prosthetic valves in vitro. A manifestation of localized gradients and pressure recovery. Circulation. 1990;82:1467–1475.

6. Niederberger J, Schima H, Maurer G and Baumgartner H. Importance of pressure recovery for the assessment of aortic stenosis by Doppler ultrasound. Role of aortic size, aortic valve area, and direction of the stenotic jet in vitro. Circulation. 1996;94:1934–40.

7. Bech-Hanssen O, Caidahl K, Wallentin I, Brandberg J, Wranne B and Ask P. Aortic prosthetic valve design and size: relation to Doppler echocardiographic findings and pressure recovery-an in vitro study. J Am Soc Echocardiogr. 2000;13:39–50.

8. Bech-Hanssen O, Gjertsson P, Houltz E, Wranne B, Ask P, Loyd D, et al. Net pressure gradients in aortic prosthetic valves can be estimated by Doppler. J Am Soc Echocardiogr. 2003;16:858–66.

9. Gjertsson P, Caidahl K, Svensson G, Wallentin I and Bech-Hanssen O. Important pressure recovery in patients with aortic stenosis and high Doppler gradients. The Am J Cardiol. 2001;88:139–44.

10. Garcia D, Pibarot P, Dumesnil JG, Sakr F and Durand LG. Assessment of aortic valve stenosis severity: A new index based on the energy loss concept. Circulation. 2000;101:765–71.

11. Bahlmann E, Cramariuc D, Gerdts E, Gohlke-Baerwolf C, Nienaber CA, Eriksen E, Wachtell, et al. Impact of pressure recovery on echocardiographic assessment of asymptomatic aortic stenosis: a SEAS substudy. JACC Cardiovasc Imaging. 2010;3:555–62.

12. Heo R, Jin X, Oh JK, Kim YJ, Park SJ, Park SW, et al. Clinical Usefulness of Pressure Recovery Adjustment in Patients with Predominantly Severe Aortic Stenosis: Asian Valve Registry Data. J Am Soc Echocardiogr. 2020;33:332–341.e2.

13. Bahlmann E, Gerdts E, Cramariuc D, Gohlke-Baerwolf C, Nienaber CA, Wachtell K, et al. Prognostic value of energy loss index in asymptomatic aortic stenosis. Circulation. 2013;127:1149–56.

14. Holy EW, Nguyen-Kim TDL, Hoffelner L, Stocker D, Stadler T, Stähli BE, et al. Multimodality imaging derived energy loss index and outcome after transcatheter aortic valve replacement. Eur Heart J Cardiovasc Imaging. 2020;21:1092–1102.

15. Strange G, Celermajer DS, Marwick T, Prior D, Ilton M, Codde J, et al. The National Echocardiography Database Australia (NEDA): Rationale and methodology. Am Heart J. 2018;204:186–189.

16. Magliano D, Liew D, Pater H, Kirby A, Hunt D, Simes J, et al. Accuracy of the Australian National Death Index: comparison with adjudicated fatal outcomes among Australian participants in the Long-term Intervention with Pravastatin in Ischaemic Disease (LIPID) study. Aust N Z J Public Health. 2003;27:649–53.

17. Playford D, Strange G, Celermajer DS, Evans G, Scalia GM, Stewart S, et al. Diastolic dysfunction and mortality in 436 360 men and women: the National Echo Database Australia (NEDA). Eur Heart J Cardiovasc Imaging. 2021;22:505–515.

18. Strange G, Stewart S, Celermajer D, Prior D, Scalia GM, Marwick T, et al. Poor Long-Term Survival in Patients With Moderate Aortic Stenosis. J Am Coll Cardiol. 2019;74:1851–1863.

19. Strange G, Stewart S, Celermajer DS, Prior D, Scalia GM, Marwick TH, et al. Threshold of Pulmonary Hypertension Associated With Increased Mortality. J Am Coll Cardiol. 2019;73:2660–2672.

20. Lang RM, Badano LP, Mor-Avi V, Afilalo J, Armstrong A, Ernande L, et al. Recommendations for cardiac chamber quantification by echocardiography in adults: an update from the American Society of Echocardiography and the European Association of Cardiovascular Imaging. European heart journal Cardiovascular Imaging. 2015;16:233–70.

21. Nagueh SF, Smiseth OA, Appleton CP, Byrd BF, 3rd, Dokainish H, Edvardsen T, et al. Recommendations for the Evaluation of Left Ventricular Diastolic Function by Echocardiography: An Update from the American Society of Echocardiography and the European Association of Cardiovascular Imaging. European heart journal Cardiovascular Imaging. 2016;17:1321–1360.

22. Maréchaux S, Ringle A, Rusinaru D, Debry N, Bohbot Y and Tribouilloy C. Prognostic Value of Aortic Valve Area by Doppler Echocardiography in Patients With Severe Asymptomatic Aortic Stenosis. J Am Heart Assoc. 2016;5.

23. Yoshida H, Seo Y, Ishizu T, Izumo M, Akashi YJ, Yamashita E, et al. Prognostic Value of Energy Loss Coefficient for Predicting Asymptomatic Aortic Stenosis Outcomes: Direct Comparison With Aortic Valve Area. J Am Soc Echocardiogr. 2019;32:351–358.e3.

24. Saeed S, Senior R, Chahal NS, Lønnebakken MT, Chambers JB, Bahlmann E, et al. Lower Transaortic Flow Rate Is Associated With Increased Mortality in Aortic Valve Stenosis. JACC Cardiovascular imaging. 2017;10:912–920.

25. Saeed S, Vamvakidou A, Zidros S, Papasozomenos G, Lysne V, Khattar RS, et al. Sex differences in transaortic flow rate and association with all-cause mortality in patients with severe aortic stenosis. Eur Heart J Cardiovasc Imaging. 2021;22:977–982.

26. Vamvakidou A, Jin W, Danylenko O, Chahal N, Khattar R and Senior R. Low Transvalvular Flow Rate Predicts Mortality in Patients With Low-Gradient Aortic Stenosis Following Aortic Valve Intervention. JACC Cardiovasc Imaging. 2019;12:1715–1724.

27. Dyverfeldt P, Hope MD, Tseng EE and Saloner D. Magnetic resonance measurement of turbulent kinetic energy for the estimation of irreversible pressure loss in aortic stenosis. JACC Cardiovasc Imaging. 2013;6:64–71.

28. Ha H, Hwang D, Kim GB, Kweon J, Lee SJ, Baek J, et al. Estimation of turbulent kinetic energy using 4D phase-contrast MRI: Effect of scan parameters and target vessel size. Magn Reson Imaging. 2016;34:715–723.

29. Delesalle G, Bohbot Y, Rusinaru D, Delpierre Q, Maréchaux S and Tribouilloy C. Characteristics and Prognosis of Patients With Moderate Aortic Stenosis and Preserved Left Ventricular Ejection Fraction. J Am Heart Assoc. 2019;8:e011036.

30. Kanamori N, Taniguchi T, Morimoto T, Watanabe H, Shiomi H, Ando K, et al. Prognostic Impact of Aortic Valve Area in Conservatively Managed Patients With Asymptomatic Severe Aortic Stenosis With Preserved Ejection Fraction. J Am Heart Assoc. 2019;8:e010198.

31. Dweck MR, Joshi S, Murigu T, Gulati A, Alpendurada F, Jabbour A, et al. Left ventricular remodeling and hypertrophy in patients with aortic stenosis: insights from cardiovascular magnetic resonance. J Cardiovasc Magn Res. 2012;14:50.

32. Spitzer E, Hahn RT, Pibarot P, de Vries T, Bax JJ, Leon MB, et al. Aortic Stenosis and Heart Failure: Disease Ascertainment and Statistical Considerations for Clinical Trials. Card Fail Rev. 2019;5:99–105.

33. Penalver J, Ambrosino M, Jeon HD, Agrawal A, Kanjanahattakij N, Pitteloud M, et al. Transthyretin Cardiac Amyloidosis and Aortic Stenosis: Connection and Therapeutic Implications. Curr Cardiol Rev. 2020;16:221–230.

